# Bayesian network modelling to identify on-ramps to childhood obesity

**DOI:** 10.1101/2022.11.23.22282647

**Authors:** Wanchuang Zhu, Roman Marchant, Richard W Morris, Louise A Baur, Stephen J Simpson, Sally Cripps

## Abstract

**Background:** When tackling complex public health challenges such as childhood obesity, interventions focused on immediate causes, such as poor diet and physical inactivity, have had limited success, largely because upstream root causes remain unresolved. A priority is to develop new modelling frameworks to infer the causal structure of complex chronic disease networks, allowing disease “on-ramps” to be identified and targeted.

**Methods:** The system surrounding childhood obesity was modelled as a Bayesian Network, using data from The Longitudinal Study of Australian Children. The existence and direction of the dependencies between factors represent possible causal pathways for childhood obesity and were encoded in directed acyclic graphs (DAGs). The posterior distribution of the DAGs was estimated using Partition Markov chain Monte Carlo.

**Results:** We have implemented structure learning for each dataset. For each wave and cohort, socio-economic status was central to the DAGs, implying that socio-economic status drives the system regarding childhood obesity. Furthermore, the causal pathway socio-economic status and/or parental high school levels → parental body mass index (BMI) → child’s BMI existed in over 99.99% of posterior DAG samples across all waves and cohorts. For children under the age of 8y, the most influential proximate causal factors explaining child BMI were birth weight and parents’ BMI. After age 8y, free time activity became an important driver of obesity, while the upstream factors influencing free time activity for boys compared with girls were different.

**Conclusions:** Childhood obesity is largely a function of socio-economic status, which is manifest through numerous downstream factors. Parental high school levels entangle with socio-economic status, hence are on-ramp to childhood obesity. The strong and independent causal relationship between birth weight and childhood BMI suggests a biological link. Our study implies that interventions that improve socio-economic status, including through increasing high school completion rates, may be effective in reducing childhood obesity prevalence.

## Introduction

Chronic diseases emerge as the outcome of complex interactions among many variables, spanning individual biology (genetics, epigenetics, metabolism, physiology, behaviours) through to environmental, social and psychological, societal, and global influences.^1^ Knowledge of this complexity has been important in moving beyond simple linear regression approaches to the prevention and treatment of chronic diseases. However, the challenge remains to tame the complexity of chronic disease systems by 1) simplifying the system, and 2) identifying key causal pathways among the tangle of influences, which can then be targeted through public health and clinical interventions.^2^

One advance towards simplifying the system has been the discovery that many chronic conditions (e.g., obesity, cardiometabolic diseases, many cancers, dementia, autoimmune diseases), as well as the biology of ageing, share a common immuno-metabolic substrate, which is powerfully modulated by diet, sleep, physical activity and mental health.^3,4^ Identifying such common mechanisms and causal structures simplifies the complex disease system, potentially rendering it more tractable to interventions that yield multiple simultaneous benefits.

When developing effective intervention targets within a complex system, it is important to distinguish immediate causal factors from influences which serve as “on-ramps” to increased risk of disease. Commonly, health interventions target immediate causes, such as poor diet or physical inactivity in the case of obesity, while leaving upstream root causes untouched and the problem unsolved.^5^ Hence, a priority is to develop modelling frameworks which can infer the causal structure of chronic disease networks.

Here we implement one of the latest techniques in causal modelling, Bayesian networks (BN), to conduct a probabilistic causal analysis of the factors leading to childhood obesity, using data from a population study of Australian children. This method has the advantage of separating causal factors into those that are immediate factors, and therefore directly connected to the outcome, from those that serve as on-ramps, and are connected indirectly via intermediate variables.^6^ Inference in BN has two parts: inference regarding the parameters of a particular network structure, and inference regarding the actual structure itself. BN studies in health care (reviewed by McLachlan et al.^7^) have largely ignored inference regarding the network structure and either assumed a particular structure *a priori* or sought the most likely structure without considering the relative probabilities of all possible structures. The latter is especially problematic when there are many near equally likely structures, as is inevitably the case within complex networks of interacting variables such as for chronic disease. To address these problems, we used a technique, known as Partition Markov chain Monte Carlo (PMCMC),^8^ to place probabilities on all possible network structures rather than selecting a single most likely network structure.

## Methods

### Data sources

Data for the analyses came from ‘Growing Up in Australia: The Longitudinal Study of Australian Children’ (LSAC),^9^ Australia’s nationally representative children’s longitudinal study, focusing on social, economic, physical, and cultural impacts on health, learning, social and cognitive development. The study tracks two cohorts of children, referred to as the birth (B) cohort (5107 infants from 0-1 years old) and the kindergarten (K) cohort (4983 children from ages 4-5 years). Data were collected over seven biennial visits (“Waves”) from 2004 to 2016.

A selection of ∼25 variables (Table 1) was chosen from the questionnaires for inclusion in Bayesian network models, informed by the existing literature on childhood obesity; e.g. the literature indicates that parental body mass index (BMI), socio-economic status, birthweight score and screen time are causally associated with childhood BMI.

**Table 1.**
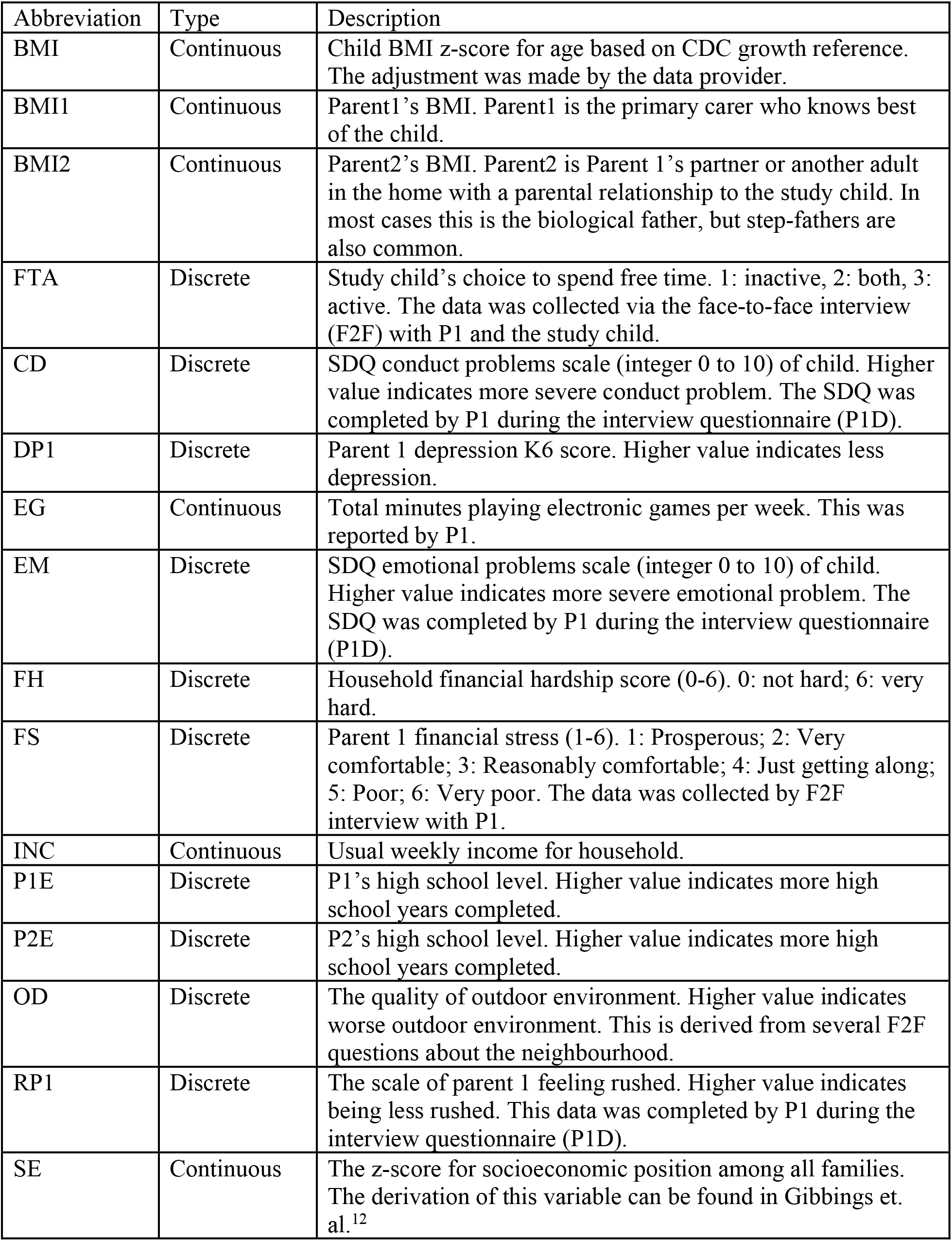
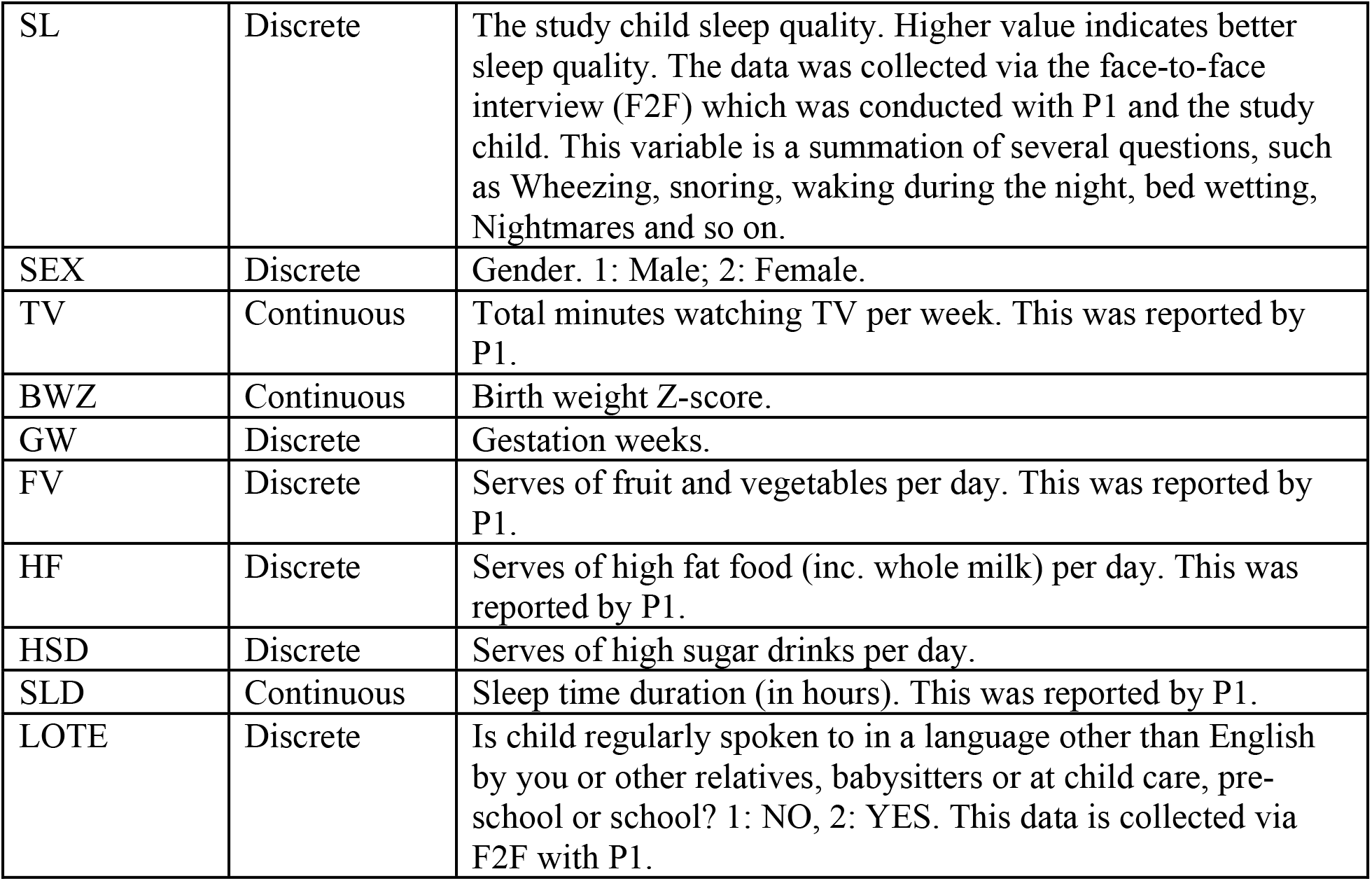
The descriptions of the variables in the analysis.

| Abbreviation | Type | Description |
| --- | --- | --- |
| BMI | Continuous | Child BMI z-score for age based on CDC growth reference. The adjustment was made by the data provider. |
| BMI1 | Continuous | Parent1's BMI. Parent1 is the primary carer who knows best of the child. |
| BMI2 | Continuous | Parent2's BMI. Parent2 is Parent 1's partner or another adult in the home with a parental relationship to the study child. In most cases this is the biological father, but step-fathers are also common. |
| FTA | Discrete | Study child's choice to spend free time. 1: inactive, 2: both, 3: active. The data was collected via the face-to-face interview (F2F) with P1 and the study child. |
| CD | Discrete | SDQ conduct problems scale (integer 0 to 10) of child. Higher value indicates more severe conduct problem. The SDQ was completed by P1 during the interview questionnaire (P1D). |
| DP1 | Discrete | Parent 1 depression K6 score. Higher value indicates less depression. |
| EG | Continuous | Total minutes playing electronic games per week. This was reported by P1. |
| EM | Discrete | SDQ emotional problems scale (integer 0 to 10) of child. Higher value indicates more severe emotional problem. The SDQ was completed by P1 during the interview questionnaire (P1D). |
| FH | Discrete | Household financial hardship score (0-6). 0: not hard; 6: very hard. |
| FS | Discrete | Parent 1 financial stress (1-6). 1: Prosperous; 2: Very comfortable; 3: Reasonably comfortable; 4: Just getting along; 5: Poor; 6: Very poor. The data was collected by F2F interview with P1. |
| INC | Continuous | Usual weekly income for household. |
| P1E | Discrete | P1's high school level. Higher value indicates more high school years completed. |
| P2E | Discrete | P2's high school level. Higher value indicates more high school years completed. |
| OD | Discrete | The quality of outdoor environment. Higher value indicates worse outdoor environment. This is derived from several F2F questions about the neighbourhood. |
| RP1 | Discrete | The scale of parent 1 feeling rushed. Higher value indicates being less rushed. This data was completed by P1 during the interview questionnaire (P1D). |
| SE | Continuous | The z-score for socioeconomic position among all families. The derivation of this variable can be found in Gibbings et. al.<sup>12</sup> |
| SL | Discrete | The study child sleep quality. Higher value indicates better sleep quality. The data was collected via the face-to-face interview (F2F) which was conducted with P1 and the study child. This variable is a summation of several questions, such as Wheezing, snoring, waking during the night, bed wetting, Nightmares and so on. |
| SEX | Discrete | Gender. 1: Male; 2: Female. |
| TV | Continuous | Total minutes watching TV per week. This was reported by P1. |
| BWZ | Continuous | Birth weight Z-score. |
| GW | Discrete | Gestation weeks. |
| FV | Discrete | Serves of fruit and vegetables per day. This was reported by P1. |
| HF | Discrete | Serves of high fat food (inc. whole milk) per day. This was reported by P1. |
| HSD | Discrete | Serves of high sugar drinks per day. |
| SLD | Continuous | Sleep time duration (in hours). This was reported by P1. |
| LOTE | Discrete | Is child regularly spoken to in a language other than English by you or other relatives, babysitters or at child care, pre-school or school? 1: NO, 2: YES. This data is collected via F2F with P1. |

### Study design

We analysed 12 of the cross-sectional datasets (Waves 2-7 in B cohort and Waves 1-6 in K cohort). For each Wave and cohort, a Bayesian network (BN)^6^ was used to model the factors surrounding childhood BMI. At each time point (Wave) the cross-sectional dataset was used to construct the distribution of possible network structures, allowing for inference on the causal pathways to childhood BMI at that time point. By comparing cross-sectional networks, we could then follow the evolution of these causal pathways over time.

To investigate the causal factors of childhood BMI in different genders, we further split each data set into boys and girls and made inferences on the corresponding Bayesian networks separately.

### Learning a Bayesian Network

When aiming to infer causality, graph structures are sought which do not contain any cycles/loops (such loops lead to self-causality, which is hard to interpret). These structures are called directed acyclic graphs (DAGs). Figure 1a illustrates a hypothetical DAG containing four variables: socio-economic status, BMI of the primary caregiver (BMI1), BMI of the second parent (BMI2), and BMI of the child (BMI). The interpretation of this DAG is as follows: First, socio-economic status is antecedent to parents’ BMI, i.e., socio-economic status is causal to the parents’ BMI and not the other way around. Second, both caregivers’ BMIs are causal to the child’s BMI. Third, conditional on the caregivers’ BMIs, a child’s BMI is independent of socio-economic status, i.e., socio-economic status has no impact on child BMI, given the parents’ BMI.

**Figure 1 legend.**
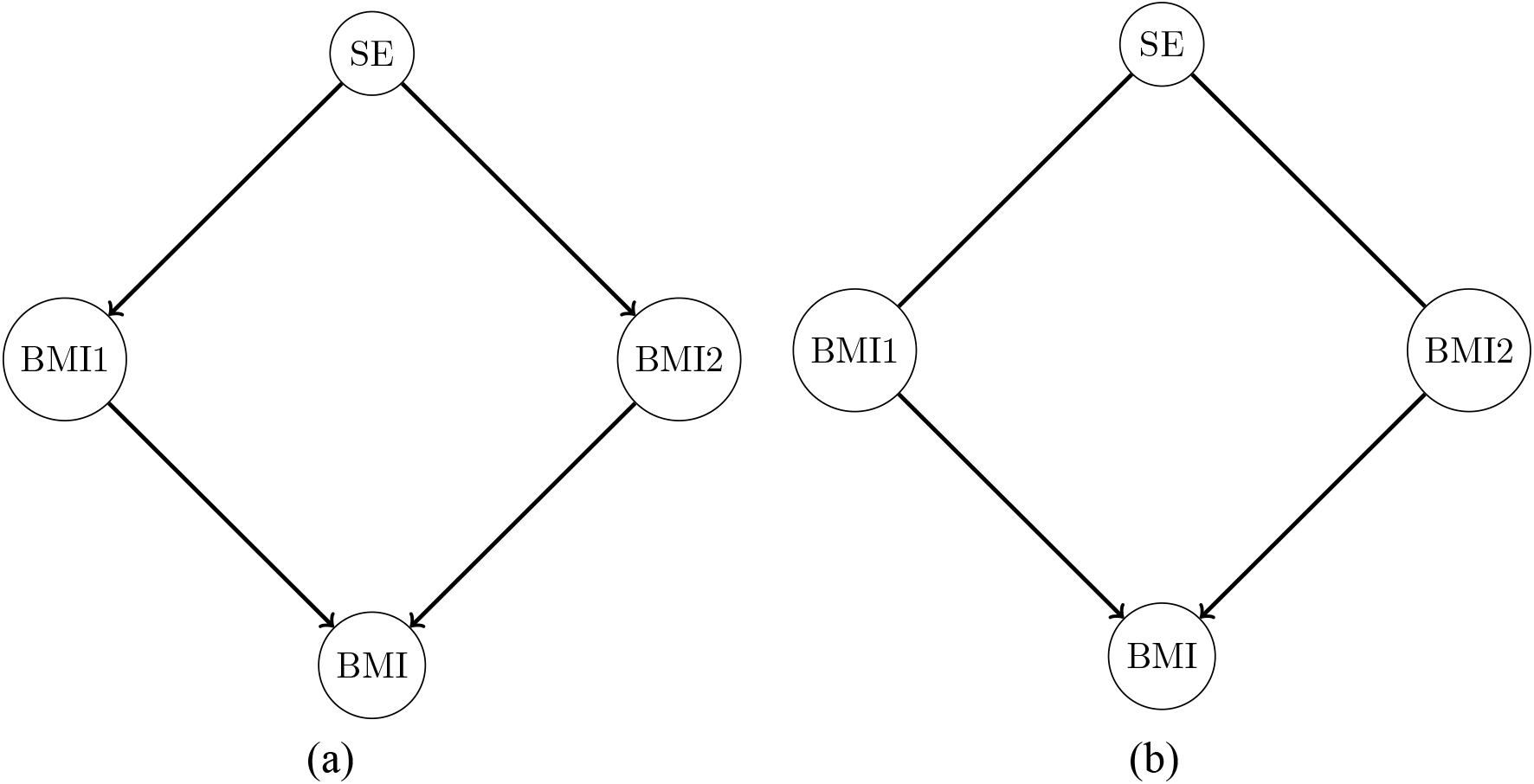
An example of directed acyclic graph (DAG) containing four nodes. A directed edge between two nodes may indicate causal relationship. For instance, SE → BMI1 could be interpreted that SE impacts BMI1. SE denotes socio-economic status, BMI1 denotes the primary caregiver’s BMI, BMI2 denotes the second caregiver’s BMI and BMI denotes the child’s BMI.

A BN can be thought of as a graphical representation of a structural equation model (SEM). In a Bayesian paradigm, one starts with a prior belief about the subject of interest (here, the DAG structure) based on existing knowledge. Then, on observing data, this prior belief is updated via what is known as a ‘likelihood function’ to arrive at a revised (‘posterior’) belief. In the context of BNs, the subject of interest has two components: first, the parameters of a particular DAG configuration, which we denote generically by *θ*_*G*_, including quantities such as the strength of the connection between two factors; and second, the DAG itself, denoted by *G*. We wish to infer both *θ*_*G*_ and *G*, which is done via the joint posterior distribution *P*(*θ*_*G*_, *G* | *data*) = *P*(*θ*_*G*_ | *G, data*)*P*(*G* | *data*). We first make inference regarding the structure *G*, by attaching probabilities to structures, *P*(*G* | *data*) and then, given a structure, infer the parameters needed to prescribe that structure *P*(*θ*_*G*_ | *G, data*). In the first step, *P*(*G* | *data*) is computed by integrating over all the possible values of parameters. This is different from traditional SEM which either assumes *G* is known or selects a single *G, Ĝ* say, using a model selection technique and then makes inference only about *θ*_*Ĝ*_.^10,11^ However, structure learning is arguably more fundamental to causal inference than parameter estimation, since the parameters can only be estimated once the structure is known.

The review by McLachlan and colleagues^7^ refers to three approaches for estimating a BN structure: data-driven, expert knowledge-driven, and hybrid approaches. These approaches are all Bayesian, which correspond to varying prior beliefs. The solely data-driven approach is analogous to a prior belief which assumes that each possible DAG is equally likely. The expert approach is analogous to a prior belief which assumes that the expert-constructed network is the true network, with probability 1.The hybrid approach, as used here, allows the strength of prior beliefs to vary both within and across structures; hence, information from different sources can be incorporated in a logically consistent manner, allowing the relative contributions of information from experts and from data to be measured. Importantly, hybrid approaches provide an ideal platform for formalising the collaboration between subject domain experts and specialist data experts: both groups are essential for success.

Although Bayesian networks have the potential to implement causal inference using observational data, they are not without drawbacks. First, the number of possible DAGs grows super-exponentially with respect to the number of variables, and it is computationally infeasible to compute the likelihood for each possible DAG once there are more than only a moderate number (∼10) of variables. Second, the structure learning algorithms can only learn up to a DAG’s equivalence class, in which all the DAGs are equally likely.^6^ The equivalence class is represented by a completed partially directed acyclic graph (CPDAG).^6^ CPDAGs contain undirected links which could be in either direction. Figure 1b shows the CPDAG of the DAG in Figure 1a. In Figure 1b, the undirected link between socio-economic status and BMI1 indicates we cannot distinguish the causal directions. For computational reasons, all the existing algorithms to estimate network structures assume that continuous variables cannot be ‘parents’ of discrete variables.^12^ In our data, there are both discrete and continuous variables. The algorithm we used to conduct structure learning is Partition Markov chain Monte Carlo (PMCMC)^7^ and the code is available at the Comprehensive R Archive Network (https://cran.r-project.org/web/packages/BiDAG/index.html). All the analyses in this paper were undertaken in R 4.0.4 (https://www.R-project.org/). PMCMC reduces the abovementioned computational challenges by collapsing the DAG space into partition space. We have adopted a strategy which considers every variable to be a Gaussian random variable to tackle the challenge caused by the existence of a mixture of continuous and discrete random variables in the data. The details can be found in the Supplementary Material.

By applying PMCMC to the LSAC data, we obtained posterior samples of DAG structures at each time point for each Wave and cohort of the LSAC data. Following the changes in DAG structures across waves allowed us to observe how causal patterns change as children age.

We also calculated the posterior probability of each DAG (top left corner), which describes the probability of each DAG given the data. These probabilities are expressed as a proportion of the sum of the posterior probability densities corresponding to the top 100 graphs. The larger the value, the more probable is the graph. Mathematically, the probability is defined as 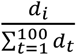, where *d*_*i*_ is the likelihood of the ith graph; i.e., a value of 70% indicates that when considering the subset of the top 100 graph structures, that graph has a posterior probability of 0.70 if each graph is equally likely *a priori*.

## Results

Table 2 lists the demographic features of the 2135 children depicted in Figure 2 (B cohort Wave 5), stratified over three weight classes according to BMI (underweight *or less*, normal weight, overweight *or greater*, based on Cole and colleagues).^13^ The pattern of mean differences between weight classes is consistent with much of the previous literature on obesity. Children with obesity were more likely to have a lower socio-economic status score and more financial hardship; were less active with more TV minutes; have parents with higher BMI; and have a higher birth weight z-score. However, these mean differences cannot elucidate the causal dependencies represented by the DAGs. See the Supplementary Material for the demographic features of the other Waves.

**Table 2.** Birth cohort aged 8 to 9

| Characteristic | Underweight<br>N = 107<sup>1</sup> | Normal<br>N = 1601<sup>1</sup> | Overweight<br>N = 427<sup>1</sup> |
| --- | --- | --- | --- |
| Female | 57 (53%) | 763 (48%) | 218 (51%) |
| BMI z-score (BMI) | -1.86 (0.68) | 0.13 (0.59) | 1.62 (0.38) |
| socioeconomic position (SE) | 0.35 (0.99) | 0.33 (0.92) | 0.06 (0.88) |
| Child's choice to spend free time (FTA) |  |  |  |
| Active | 29 (27%) | 430 (27%) | 86 (20%) |
| Active and inactive | 55 (51%) | 750 (47%) | 199 (47%) |
| Inactive | 23 (21%) | 420 (26%) | 142 (33%) |
| Total No. of TV minutes for an average week (TV) | 12 (7) | 13 (8) | 14 (8) |
| Total No. of electronic game minutes for an average | 5.3 (5.2) | 5.0 (4.9) | 5.3 (5.4) |
| SDQ Emotional symptoms scale (EM) | 2.06 (1.99) | 1.62 (1.76) | 1.82 (1.87) |
| SDQ Conduct problems scale (CD) | 1.00 (1.14) | 1.08 (1.30) | 1.30 (1.44) |
| Weekly household income (annual) (INC) |  |  |  |
| \$0-\$999 (\$0-\$51,999) | 10 (9.3%) | 86 (5.4%) | 46 (11%) |
| \$1,000-\$1,999 (\$52,000-\$103,999) | 34 (32%) | 516 (32%) | 133 (31%) |
| \$2,000-\$2,999 (\$104,000-\$155,999) | 37 (35%) | 540 (34%) | 154 (36%) |
| \$3,000 or more (\$156,000 or more) | 26 (24%) | 459 (29%) | 94 (22%) |
| How family is getting on financially (FS) |  |  |  |
| Prosperous/Very comfortable | 25 (23%) | 533 (33%) | 113 (26%) |
| Comfortable/getting along | 82 (77%) | 1,057 (66%) | 308 (72%) |
| Poor/Very poor | 0 (0%) | 11 (0.7%) | 6 (1.4%) |
| Hardship scale (FH) | 0.15 (0.45) | 0.13 (0.51) | 0.21 (0.56) |
| Parental school completion (P1E) | 78 (73%) | 1,281 (80%) | 302 (71%) |
| Parental school completion (P2E) | 68 (64%) | 1,088 (68%) | 265 (62%) |
| Parental body mass index (BMI1) | 24.2 (4.9) | 25.5 (5.1) | 28.8 (6.0) |
| Parental body mass index (BMI2) | 25.7 (3.3) | 27.3 (3.9) | 29.4 (4.7) |
| Frequency of feeling rushed (RP1) |  |  |  |
| Always/Often | 65 (61%) | 992 (62%) | 242 (57%) |
| Sometimes | 37 (35%) | 502 (31%) | 149 (35%) |
| Rarely/Never | 5 (4.7%) | 107 (6.7%) | 36 (8.4%) |
| K-6 Depression scale summed score (DP1) | 8.57 (2.65) | 8.42 (2.83) | 8.86 (3.33) |
| Frequency ate fruit and vegetables (FV) | 3.19 (1.25) | 3.41 (1.38) | 3.28 (1.38) |
| Frequency ate high fat food (inc. whole milk) (HF) | 3.20 (1.41) | 3.24 (1.43) | 3.12 (1.54) |
| Frequency drank high sugar drinks (HSD) | 0.99 (1.08) | 0.94 (1.02) | 1.04 (1.07) |
| Poor sleep quality (SL)<sup>a</sup> | 33 (31%) | 463 (29%) | 116 (27%) |
| Wake up in the morning (Time) (SLD) | 612 (43) | 618 (38) | 611 (43) |
| Child regularly spoken to in a language other than English | 23 (21%) | 251 (16%) | 69 (16%) |
| No. weeks of gestation (GW) | 38.23 (6.01) | 38.92 (3.76) | 39.01 (3.44) |
| Birth weight z-score (BWZ) | -0.42 (0.99) | 0.04 (1.04) | 0.21 (1.10) |

<sup>1</sup>n (%); Mean (SD); <sup>a</sup>Sleep problems > 0

**Fig 2 legend.**
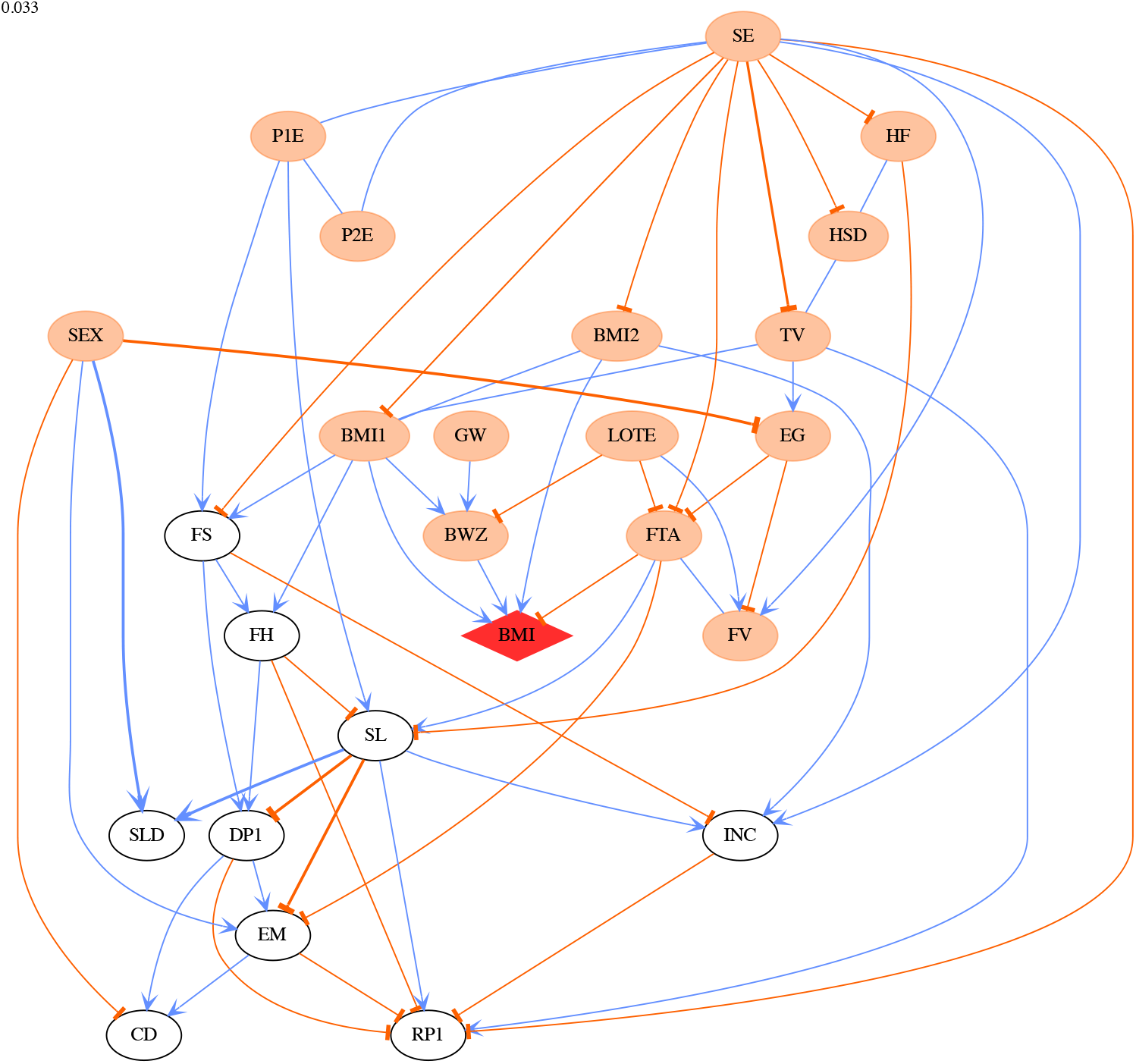
The CPDAG derived from the most probable DAG for Wave 5 in B cohort. The child BMI node is highlighted by a red diamond shape. The thicknesses of the edges in the network correspond to the strength of relationship between nodes exists, with a thicker line denoting a higher absolute value. The link and orange edges indicate positive and negative relationships respectively. The ellipse nodes in each plot are colour-coded as follows: an orange node denotes ancestors of child BMI.

### Central role of socio-economic status and parental education over all time points

The CPDAG derived from the most probable DAG for B cohort Waves 5 (age 8-9) is shown in Figure 2. It clearly shows that socio-economic status played a central role in the obesity networks we studied. For every Wave in the B cohort, socio-economic status sits in the central position of the CPDAG structure. This implies that socio-economic status drives almost everything else in the network structure. The same conclusion applies to other Waves. In LSAC, socio-economic status was derived from family income, parents’ education and parents’ occupational status (Gibbings and colleagues),^14^ however our results indicate that socio-economic status represents an important influence on child BMI over and above any of its constituents alone. In addition, more than 99% of the posterior samples of DAG structures contain a pathway from socio-economic status or parental high school level to child BMI. The detail of percentages is found in Table 3.

**Table 3.** The percentage of the path (SE/P1E/P2E → BMI1/BMI2 → BMI) appearing in the posterior samples for every Wave.

| Wave | 1 | 2 | 3 | 4 | 5 | 6 | 7 |
| --- | --- | --- | --- | --- | --- | --- | --- |
| B cohort | NA | 1.000 | 1.000 | 0.998 | 0.999 | 1.000 | 1.000 |
| K cohort | 0.996 | 1.000 | 1.000 | 1.000 | 1.000 | 1.000 | NA |

DAG structures from every Wave show the importance of both parents finishing high school (P1E for mother, P2E for father). These two variables are correlated with socio-economic status, and the relationships are present in every DAG. The importance of this relationship is especially apparent in the network for K cohort Wave 1 (Figure 3), for which no specific socio-economic status variable was available. Figure 3 shows that in the absence of a specific socio-economic variable, parental high school level becomes the central node for the network.

**Fig 3 legend.**
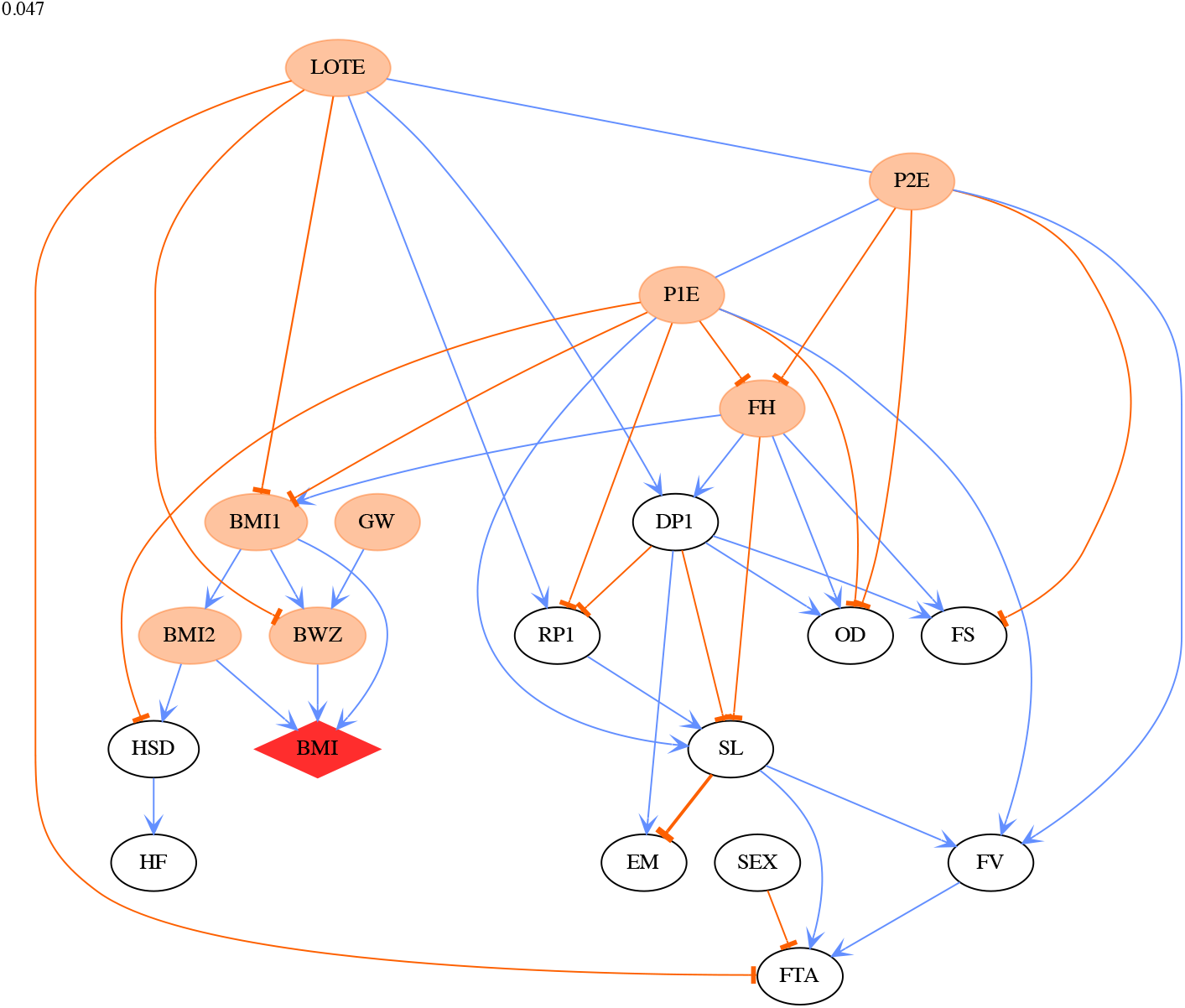
The CPDAG derived from the most probable DAG for Wave 1 in K cohort. The child BMI node is highlighted by a red diamond shape. The thicknesses of the edges in the network correspond to the strength of relationship between nodes exists, with a thicker line denoting a higher absolute value. The link and orange edges indicate positive and negative relationships respectively. The ellipse nodes in each plot are colour-coded as follows: an orange node denotes ancestors of child BMI.

### Free time activity becomes a driver of obesity as children age

For children up to the age of 6 years (Figure 4), a child’s BMI is on the periphery of the DAG and is connected to the other variables only via the BMI of the child’s carers (BMI1 and BMI2) and the child’s birth weight z-score. After the age of 6 years, the drivers of childhood obesity become more complex. There is a formation of another sub-graph around child-specific variables, such as conduct disorder, emotional problems, sleep quality and quantity and electronic games, although there is considerable uncertainty associated with the direction and strength of these relationships at different Waves.

**Fig 4 legend.**
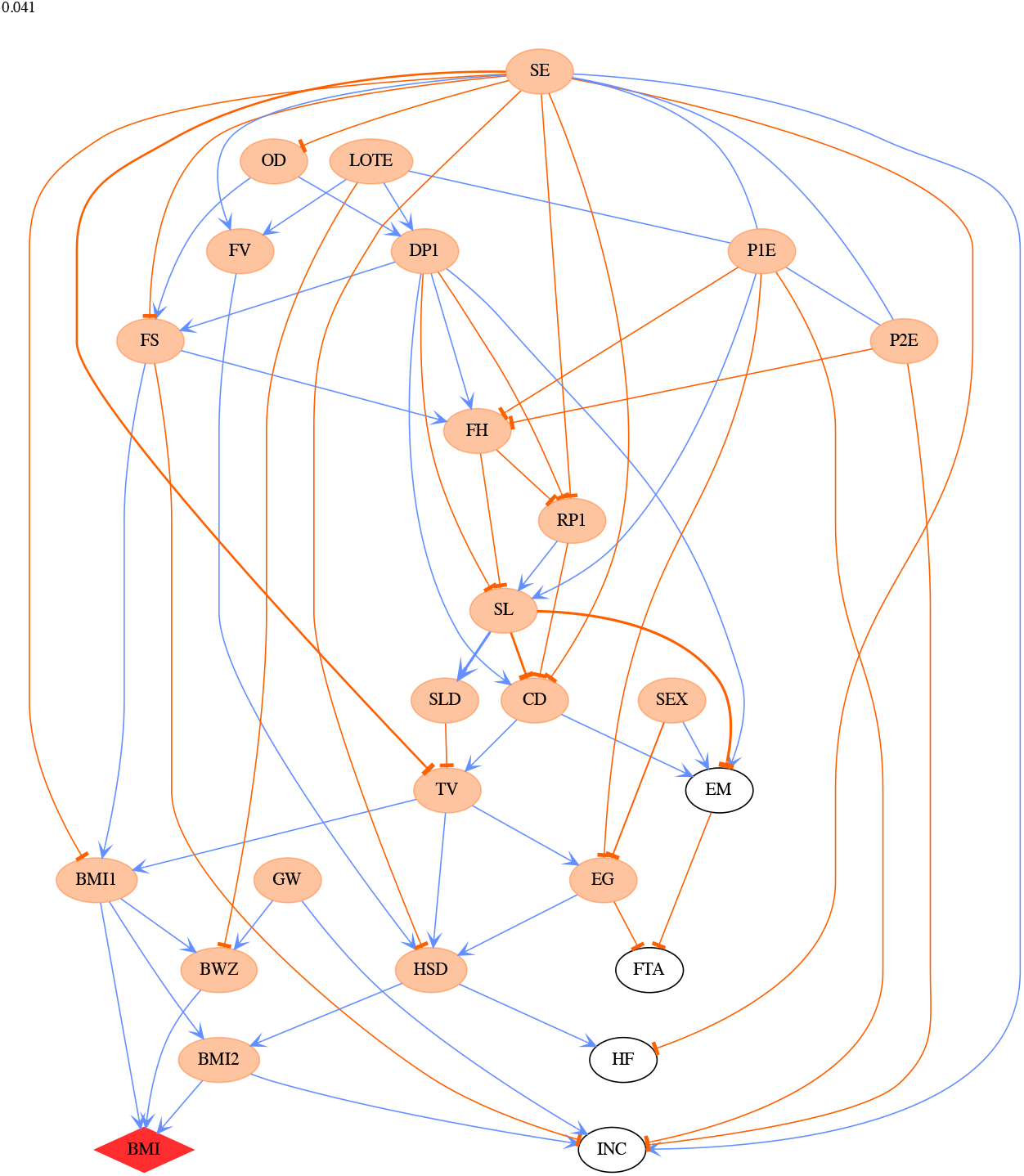
The CPDAG derived from the most probable DAG for Wave 4 in B cohort. The child BMI node is highlighted by a red diamond shape. The thicknesses of the edges in the network correspond to the strength of relationship between nodes exists, with a thicker line denoting a higher absolute value. The link and orange edges indicate positive and negative relationships respectively. The ellipse nodes in each plot are colour-coded as follows: an orange node denotes ancestors of child BMI.

Figure 2 shows that after age 8 years, free time activity (e.g., dancing and sports) becomes an important driver of obesity, and this, in turn, is driven by socio-economic status and the extent of electronic gaming by the child. Figure 4 also indicates that gender begins to impact a child’s BMI from age 6 (B cohort Wave 4). However, gender does not directly influence a child’s BMI; rather, it passes its influence through other paths, e.g., SEX → electronic gaming→ free time activity → child BMI, which is shown in Figure 2. To further investigate the impact of gender, we applied PMCMC to boys and girls separately. The CPDAG derived from the most likely DAG of B cohort Wave 5 is presented in Figure 5 for boys (Figure 5a) and girls (Figure 5b), respectively. For boys, the causal pathway electronic gaming → free time activity → child BMI emerges. However, for girls, sleep → free time activity → child BMI is the main pathway regarding how free time activity impacts child BMI. It would appear that boys and girls have different upstream factors influencing free time activity.

**Fig 5 legend.**
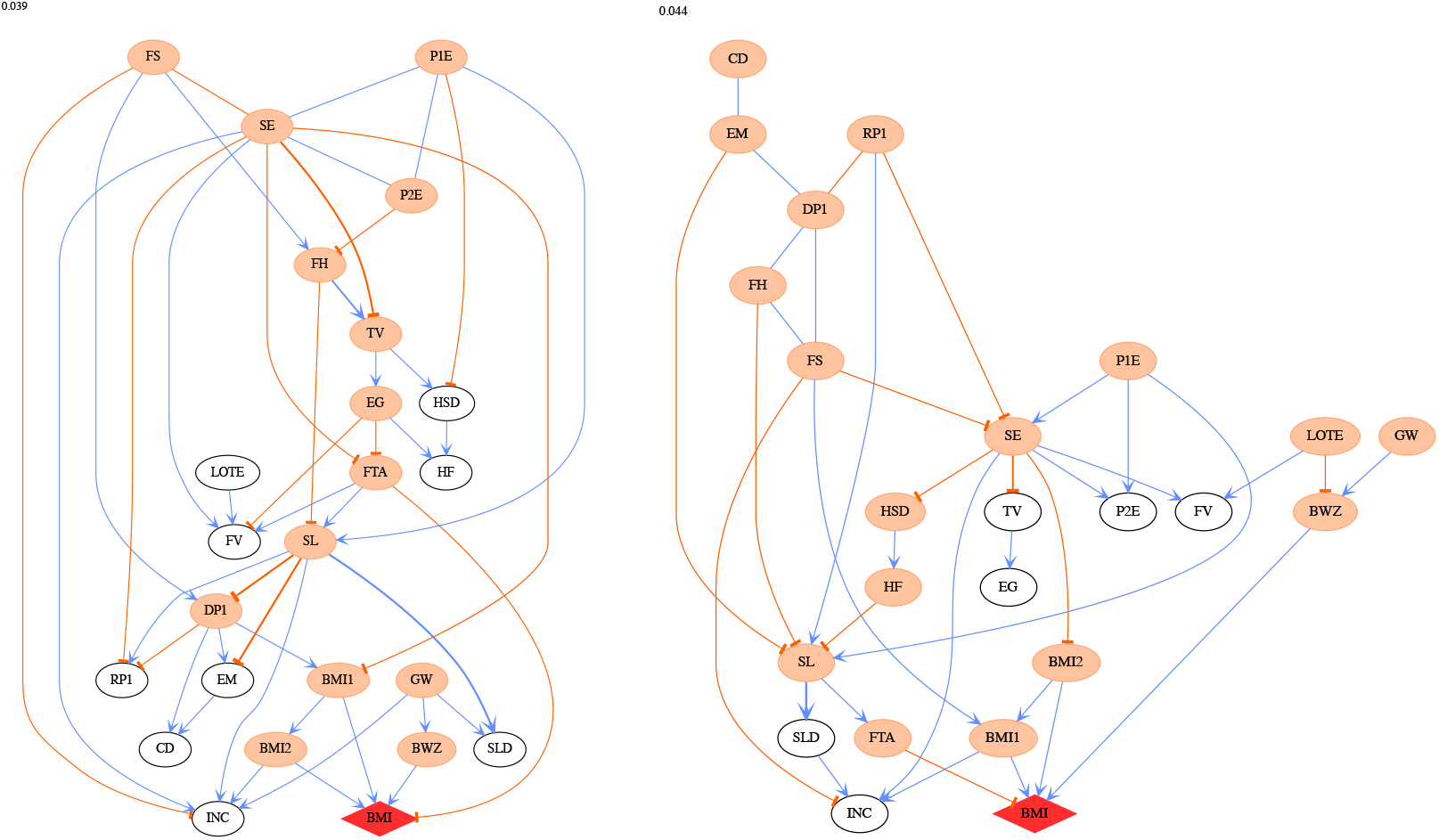
The CPDAG derived from the most probable DAG for boys and girls respectively in Wave 5 B cohort. The child BMI node is highlighted by a red diamond shape. The thicknesses of the edges in the network correspond to the strength of relationship between nodes exists, with a thicker line denoting a higher absolute value. The link and orange edges indicate positive and negative relationships respectively. The ellipse nodes in each plot are colour-coded as follows: an orange node denotes ancestors of child BMI.

To illustrate the difference between BN and multiple regression, we conducted analyses using both techniques on a dataset containing variables: child BMI, parents’ BMI, socio-economic status, and parental high school level. Child BMI was the dependent variable in multiple regression analysis, and we compared its results to that of BN. The most probable DAG obtained by PMCMC showed the complete set of direct and indirect causal pathways from each of the variables to child BMI. However, multiple regression only revealed the direct paths between parental BMIs and children’s BMI, with the other indirect relationships not detected. More details of this comparison can be found in the Supplementary Material.

## Discussion

We have used BN to infer the causal pathways leading to childhood obesity and shown how this pathway changes as children age. Our analysis of the LSAC data demonstrated that parental high school level (both paternal and maternal) serves as an on-ramp to childhood obesity. When children were aged 2-4 years the causal pathway was: socio-economic status/parental high school level → parental BMI → child BMI. By the time the child was 8-10 years old, an additional pathway had emerged: parental high school level − socio-economic status → electronic games → free time activity → child BMI.

Obesity is a complex health issue, with multiple factors that operate at the level of the individual, family and beyond contributing to its development and maintenance.^1,15,16^ For example, strong positive associations between parental and offspring BMI have been documented in many studies using traditional regression analytic approaches.^17–19^ A range of other individual, family and socio-demographic characteristics are also associated with childhood obesity, including poor dietary intake, lower levels of physical activity, higher recreational screen time, family income and parental high school levels.^18,20,21^ Studies in high income countries have shown that social disadvantage, measured via family or parental income, parental high school level, occupation or employment status, is associated in childhood with both higher obesity prevalence rates and a range of obesity-related behaviours.^18,22^

Such complexity has made it challenging to identify key causal pathways and hence to implement effective interventions.^23^ Our analyses have not only reinforced previous findings in relation to the multiple factors associated with childhood obesity but have now clarified the causal structure that underpins these associations. We have highlighted the central role of lower socio-economic status and low high school level for parents as the primary root cause of childhood obesity, which exerts its effect via several more proximal factors. Among these downstream factors, there was a strong and independent positive relationship between birth weight and childhood obesity, in keeping with findings from studies using traditional regression analyses.^24^ Birth weight itself is influenced by a range of genetic, epigenetic, maternal, in utero and social factors.

It is this ability to infer complex causal structures without temporal information which makes BN such a powerful and useful technique in health and medical research. Causal inference is achieved by estimating the full joint distribution of potential factors as a product of conditionally independent distributions, thereby distinguishing between direct and indirect dependencies. In contrast, more conventional multiple regression techniques lack a mechanism to infer causality without temporal information.^25^ Indeed, multiple regression can be considered a specific example of a BN, where a particular dependency structure is imposed *a priori*, namely that all independent variables are directly related to the dependent variable. The marked difference between these two approaches is illustrated in the two distinct causal pathways shown in the Supplementary Materials, developed using a cut-down version of our dataset.

In contrast to the structural equation modelling (SEM), another popular causal model, Bayesian networks *learn* the causal links, and the corresponding probabilities from the data, while SEM requires users either to specify the causal model prior to parameter estimation, based on expert knowledge or select an optimal structure based on some model selection criteria. ^10,11^ In our analysis, the computational challenge is greatly alleviated, firstly, by working closely with content experts to incorporate domain knowledge by constructing a form of “blacklist” in DAG structures, which includes all forbidden links, i.e., those considered by domain experts to be illogical or infeasible (see Supplementary Materials for full “blacklist”). Secondly, PMCMC is used to reduce the DAG space by grouping individual DAG structures into partitions.^8^ Importantly, PMCMC also allows samples to be drawn from the posterior distribution over graphs and thereby to quantify uncertainty, which is of paramount importance for domain practitioners who use the resulting graph structures to make decisions.

Our results have important implications for interventions to address the complex issue of childhood obesity and demonstrate why intervening at the level of more proximate, downstream factors risks leaving the root causes of childhood obesity untouched, leaves the problem unsolved. It is well recognised that low levels of maternal and paternal high school levels are associated with inequalities in child health status and mortality.^26,27^ These disparities appear to be mediated through other social determinants of health, including socio-economic status and living conditions.^28^ There is some evidence that interventions which improve parental, especially maternal, education are associated with improvements in general measures of early childhood health and child mortality^29^.However, to our knowledge there have been no such studies that measure offspring weight status by mid-childhood or adolescence. Our analyses imply that interventions that improve socio-economic status, including through increasing high school completion rates, may lead to improvements in childhood obesity prevalence over much longer time spans.

### Limitations

The LSAC data were collected in Australia which is a developed country. Thus, the children in this data set may only be representative of wealthy countries. It does not necessarily cover the characteristics of children from low- and middle-income countries.

Our study used Bayesian networks to model the variables surrounding childhood obesity. Whereas BNs are powerful, they are not without their drawbacks. They are computationally expensive, due to the super-exponential growth of the number of possible graph structures. For example, a system with 20 factors has an order of 2^190^ possible graph structures, which is greater than the number of atoms in the universe. Therefore, exhaustive search is impossible and some constraints on the number of possible graph structures need to be imposed.

All the presented causal pathways are only valid for the LSAC data. There is the possibility that some confounders were not measured in these data and misleading causal links may have resulted. For example, there could be further ‘upstream’ variables influencing both socio-economic status and parental high school levels which might explain the apparent undirected link between those two variables. However, under the current dataset, socio-economic status and parental high school levels are co-dependent.

## Data Availability

All data produced are available online at https://growingupinaustralia.gov.au/data-and-documentation/accessing-lsac-data

## Authors’ contributions

WZ (guarantor) led the study and was involved in conception, design, management, analysis, and prepared the final draft. RM was lead data scientist and was involved in conception, design, management, and wrote sections of the final draft and edited it. RWM led the demographical analysis of the data and was involved in analysis, and edited the final draft. LAB led the discussion on childhood obesity and was involved in the design, wrote sections of the final draft and edited it. SJS led discussion on childhood obesity, and was involved in the design and edited the final draft. SC was the senior author and was involved in design, management, and wrote and edited the final draft. The decision to submit was made by the senior author (SC), WZ, LAB and SJS, in conjunction with all of the authors. WZ, RM, RWM and SC had full access to the data. The corresponding author attests that all listed authors meet authorship criteria and that no others meeting the criteria have been omitted.

## Availability of data and materials

The LSAC data is available under request to anyone. The online application is at https://growingupinaustralia.gov.au/data-and-documentation/accessing-lsac-data. The data include deidentified participant data and data dictionary. The related document is also publicly available.

## Ethics approval and consent to participate

Not required.

## Consent for publication

Not applicable.

## Acknowledgements

We would like to express our sincere gratitude to the families in the project and parties which collected and shared the data.

## Abbreviations

DAG: directed acyclic graphs
BMI: parental body mass index
BN: Bayesian network
PMCMC: Partition Markov chain Monte Carlo
LSAC: The Longitudinal Study of Australian Children’
SEM: structural equation model
CPDAG: completed partially directed acyclic graph

Other abbreviations can be found in Table 1

