## supplementary for "Bayesian network modelling to identify on-ramps to childhood obesity"

### Supplementary Materials for ‘Bayesian network modelling to identify on-ramps to childhood obesity’

#### Data source: LSAC

In 2004, LSAC recruited a cross-sequential sample of 5107 infants aged 0-1 years, referred to as the birth (B) cohort, and a sample of 4983 children aged 4-5 years, referred to as the kindergarten (K) cohort, and collected data on these children in seven biennial visits from 2004 until 2016. One of the aims of the project is to identify policy opportunities to improve comprehensive support for children and their families, and to identify opportunities for early intervention. The data were collected using a variety of methods: parent self-complete or leave behind questionnaires; face-to-face interviews, usually with the parent who knew the child best; physical measurements of the child; and audio computer-assisted interviews, for children 10 years and older. In self-complete questionnaires, parents were asked questions regarding: the child’s personality and behaviours; the style of parenting; parents’ health and wellbeing; family and community circumstances; work and family; and family life. In face-to-face interviews, the parents were asked questions regarding: the child’s health and education; the type of childcare and family activity; and parental background, including income, housing & neighbourhood. More details about these data sets can be found at the official website of LSAC (https://growingupinaustralia.gov.au/).

##### Design of LSAC

Table S1 gives the design of LSAC data.

| **Cohort** | **Wave 1** | **Wave 2** | **Wave 3** | **Wave 4** | **Wave 5** | **Wave 6** | **Wave 7** |
|---|---|---|---|---|---|---|---|
| Year | 2004 | 2006 | 2008 | 2010 | 2012 | 2014 | 2016 |
| Infant (B) | 0-1y | 2-3y | 4-5y | 6-7y | 8-9y | 10-11y | 12-13y |
| Child (K) | 4-5y | 6-7y | 8-9y | 10-11y | 12-13y | 14-15y | 16-17y |

Table S1. Design of the LSAC data collection.

##### Data availability in LSAC

Table S2 gives details of the availability of the chosen variables by wave and cohort.

| Cohort | B | | | | | | | K | | | | | | |
|---|---|---|---|---|---|---|---|---|---|---|---|---|---|---|
| Wave | 1 | 2 | 3 | 4 | 5 | 6 | 7 | 1 | 2 | 3 | 4 | 5 | 6 | 7 |
| BMI | | | | | | | | | | | | | | |

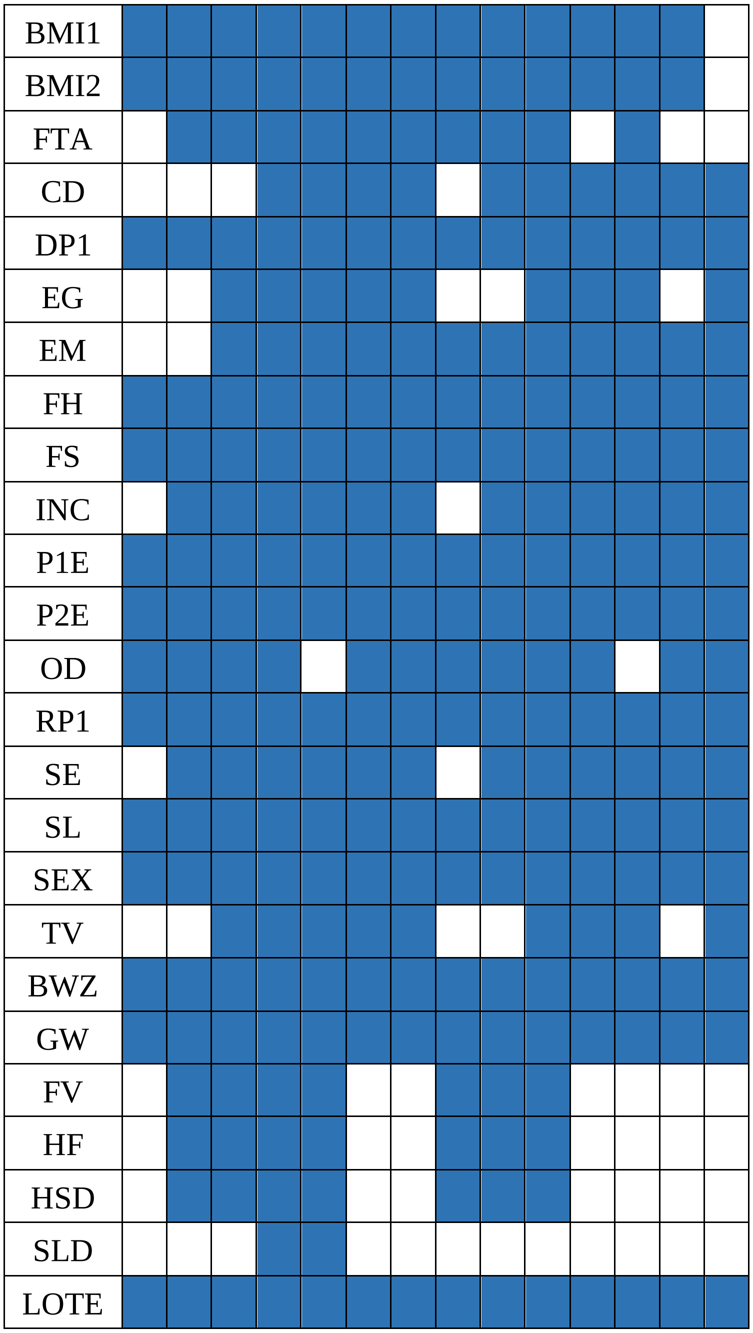


Table S2. The availability of variables in different waves for cohort B and K respectively. The white cells indicates missing values.

#### Data Pre-processing

We believe that some data entries were incorrectly entered and need to be preprocessed before modelling. For example, a child's BMI is reported as a z-score. That is, the reported value is the actual BMI minus the mean BMI and then this difference is divided through by the standard deviation of BMI. If the factor is approximately normally distributed then we would expect

99.8% of values to be in the interval [-3,3], 99.99% of values should be in the interval [-4,4], 99.9999% of values within the interval [-5,5]. Therefore, values such as -20, seem implausible, and all entries with a z-score outside the interval [-5,5] were removed. We also defined values of BMI1 or BMI2 which were greater than 60, as incorrect as a BMI of 60 seems implausible.

As a result of missing values or incorrect entries, the number of children for whom data was available was less than 5,000 and varied by cohort and by wave. Table S2 details the number of factors available for each cohort and wave.

#### Prior Setting on Structure Learning

Normally, the prior distribution on graph $G$ could be a discrete uniform distribution over the full DAG space formed by the random variables of interest. However, common sense dictates some causal links are not plausible. For example, a causal link BWZ → BMI1 does not make sense, because the primary carer's BMI exists before the child's birth weight. Table S3 lists all the implausible causal links for LSAC data. Implausible links between factors are those that violate chronological order or those links that are static, for example a child's sex will not determine a family's socio-economic status. Given the implausible links, the prior distribution of $G$ is taken to be a discrete uniform distribution over the space of DAG formed by the variables of interest, except those DAGs which contain any link described in Table S3. The abbreviations of variables names can be found in the table 1 of main text.

| **From** | **To** |
|---|---|
| BMI, GW, BWZ | BMI1 |
| BMI, GW, BWZ | BMI2 |
| FTA, SEX, BMI, TV, EG, EM, CD, FS, FH, RP1, DP1, SL, HSD, FV, HF, SLD | BWZ |
| BMI, FTA, CD, EG, EM, SEX, TV, SL, OD, GW, BWZ, FV, HF, HSD, SLD | SE |
| SEX, BMI, FTA, EG, EM, CD, DP1, FS, FH, INC, P1E, P2E, OD, SE, RP1, SL, TV, BWZ, FV, HF, HSD, SLD | GW |
| SEX, BMI, FTA, TV, EG, EM, CD, INC, FS, FH, RP1, DP1, SL, OD, GW, BWZ, HSD, SLD | P2E |
| SEX, BMI, FTA, TV, EG, EM, CD, INC, FS, FH, RP1, DP1, SL, OD, GW, BWZ, HSD, SLD | P1E |
| BMI, FTA, SE, TV, EG, EM, CD, INC, P1E, FE1, P2E, BMI1, BMI2, FS, FH, RP1, DP1, SL, OD, GW, BWZ, SLD, LOTE, FV, HF, HSD | SEX |
| BMI, FTA, BMI1, BMI2, TV, EG, EM, CD, INC, FS, FH, RP1, DP1, SL, OD, GW, BWZ, SEX, FV, HF, HSD, SLD | LOTE |
| BMI | All the others |

Table S3. The implausible directed links from prior knowledge.

##### The strategy in Partition MCMC to handle hybrid Bayesian networks

Hybrid Bayesian networks contain both discrete and continuous random variables. The specific challenge in structure learning for hybrid Bayesian networks lies in difficulty of modelling dependencies between continuous and discrete random variables. The most popular approach in the literature to this problem is to discretize the continuous variables to be discrete variables. However, the recent research shows that treating every variable as being a Gaussian distribution increases both computational efficiency and estimation accuracy.[1] Therefore, we adopt the above strategy in our analysis. That is, we treat all the discrete random variables as Gaussian random variables. Thus, Gaussian Bayesian networks were used to model our data.

**Comparison between multiple regression and Bayesian networks**

We have chosen the data in B cohort Wave 4 to illustrate the different results obtained from an analysis using multiple regression versus an analysis using Bayesian networks. Only a subset of the variables are used for this comparison. The variables include SE, P1E, P2E, BMI, BMI1 and BMI2.

Figure S1 shows the most probable DAG obtained from partition MCMC through Bayesian networks modelling. As we can see, parents' BMI are the direct causal factors to child BMI. This is consistent with the regression model, whose results are summarized in Table S4. However, Figure S1 shows more information about the structural pathways and mutual interactions. For example, Figure S1 shows that a family's SE is the centre of the network and that it strongly influences BMI1 and BMI2. Considering the equivalence classes of the DAG in Figure S1, it is concluded that P1E and P2E entangle with SE and thus jointly influence parents' BMI. Such information will not be revealed by a multiple regression analysis.

| | Estimate | Std. Error | t value | Pr(>\|t\|) |
|---|---|---|---|---|
| (Intercept) | -1.533411 | 0.246817 | -6.213 | 6.17E-10*** |
| SE | -0.025584 | 0.025606 | -0.999 | 0.318 |
| BMI1 | 0.033419 | 0.003767 | 8.872 | 2E-16*** |
| BMI2 | 0.036192 | 0.004985 | 7.259 | 5.3E-13*** |
| P1E | -0.020839 | 0.02835 | -0.735 | 0.462 |
| P2E | 0.029638 | 0.023714 | 1.25 | 0.211 |

Table S4. The estimation using linear regression.

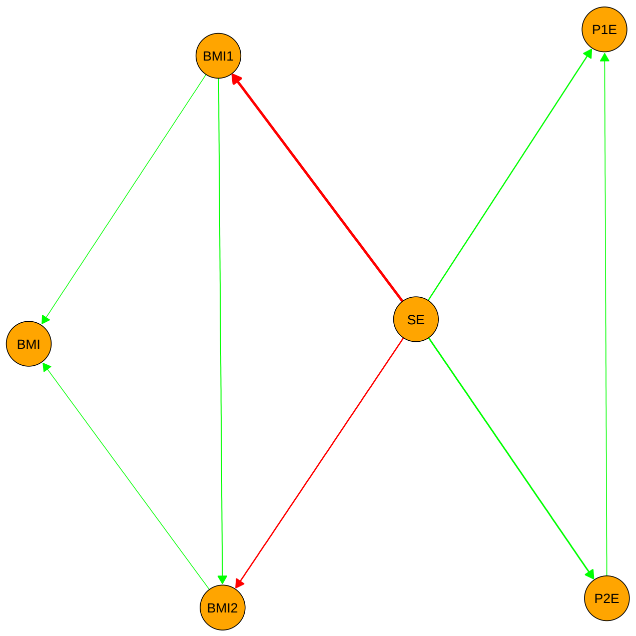


Figure S1. The most probable DAG learned by Partition MCMC.

#### Demographics for different dataset

Tables S5-S8 give demographic information broken down by three classes of childhood BMI. The categories of child BMI (underweight or less, normal weight, overweight or greater) are computed according to the criteria in Cole and colleagues.[2]

Table S5. Kindergarten cohort aged 4 to 5

| Characteristic | Underweight N = 137[1] | Normal N = 2071[1] | Overweight N = 517[1] |
|---|---|---|---|
| Female | 69 (50%) | 984 (48%) | 290 (56%) |
| BMI z-score (BMI) | -1.86 (0.84) | 0.33 (0.61) | 1.86 (0.52) |
| SC regularly spoken to in a language other then english | 29 (21%) | 358 (17%) | 94 (18%) |
| SC's choice to spend free time (FTA) | | | |
| Active | 37 (27%) | 551 (27%) | 126 (24%) |
| Active and inactive | 68 (50%) | 1,030 (50%) | 245 (47%) |
| Inactive | 32 (23%) | 490 (24%) | 146 (28%) |
| SDQ emotional symptoms scale (EM) | 1.76 (1.70) | 1.57 (1.57) | 1.53 (1.56) |
| How family is getting on financially (FS) | | | |
| Prosperous/Very comfortable | 22 (16%) | 458 (22%) | 97 (19%) |
| Comfortable/getting along | 112 (82%) | 1,568 (76%) | 409 (79%) |
| Poor/Very poor | 3 (2.2%) | 43 (2.1%) | 11 (2.1%) |
| Hardship scale (FH) | 0.33 (0.68) | 0.33 (0.75) | 0.39 (0.83) |
| Parental school completion (P1E) | 89 (65%) | 1,390 (67%) | 329 (64%) |
| Parental school completion (P2E) | 85 (62%) | 1,212 (59%) | 263 (51%) |
| Body mass index (BMI1) | 23.7 (3.8) | 24.8 (4.8) | 27.3 (6.1) |

| Characteristic | Underweight N = 137[1] | Normal N = 2071[1] | Overweight N = 517[1] |
|---|---|---|---|
| Body mass index (BMI2) | 25.6 (3.5) | 26.6 (4.0) | 27.9 (4.2) |
| Frequency of feeling rushed (RP1) | | | |
| Always/Often | 64 (47%) | 1,058 (51%) | 281 (54%) |
| Sometimes | 57 (42%) | 800 (39%) | 189 (37%) |
| Rarely/Never | 16 (12%) | 213 (10%) | 47 (9.1%) |
| K-6 Depression scale summed score (DP1) | 9.8 (3.3) | 9.8 (3.4) | 9.8 (3.3) |
| Serves of fruit and vegetables (FV) | 2.90 (1.41) | 3.03 (1.39) | 3.02 (1.41) |
| Serves of high fat food (inc. whole milk) (HF) | 3.49 (1.39) | 3.44 (1.38) | 3.36 (1.45) |
| Serves of high sugar drinks (HSD) | 1.63 (1.20) | 1.59 (1.16) | 1.70 (1.23) |
| Poor sleep quality (SL)[a] | 137 (100%) | 2,071 (100%) | 517 (100%) |
| No. weeks of gestation (GW) | 37.61 (7.35) | 38.68 (5.45) | 38.58 (5.80) |
| Birth weight z-score (BWZ) | -0.61 (1.03) | -0.05 (1.02) | 0.27 (1.14) |
| Outdoor environment quality (OD) | 1.95 (0.52) | 1.92 (0.46) | 1.96 (0.45) |

[1]n (%); Mean (SD); [a]Sleep problems > 0

#### Table S6. Birth cohort aged 6 to 7

| Characteristic | Underweight N = 114[1] | Normal N = 1825[1] | Overweight N = 422[1] |
|---|---|---|---|
| Female | 55 (48%) | 891 (49%) | 218 (52%) |
| BMI z-score (BMI) | -1.94 (0.68) | 0.20 (0.59) | 1.68 (0.41) |
| Socio-economic position (SE) | 0.42 (0.88) | 0.31 (0.91) | 0.15 (0.93) |
| Child's choice to spend free time (FTA) | | | |
| Active | 54 (47%) | 819 (45%) | 180 (43%) |
| Active and inactive | 29 (25%) | 562 (31%) | 116 (27%) |
| Inactive | 31 (27%) | 443 (24%) | 126 (30%) |
| Total No. of TV minutes for an average week (TV) | 11 (6) | 12 (7) | 14 (8) |
| Total No. of electronic game minutes for an average | 2.76 (3.39) | 2.60 (3.36) | 3.06 (4.84) |
| SDQ Emotional symptoms scale (EM) | 1.97 (2.02) | 1.57 (1.67) | 1.61 (1.77) |
| SDQ Conduct problems scale (CD) | 1.61 (1.57) | 1.35 (1.39) | 1.37 (1.40) |
| Weekly household income (annual) (INC) | | | |
| $0-$999 ($0-$51,999) | 10 (8.8%) | 174 (9.5%) | 50 (12%) |
| $1,000-$1,999 ($52,000-$103,999) | 45 (39%) | 731 (40%) | 172 (41%) |
| $2,000-$2,999 ($104,000-$155,999) | 31 (27%) | 572 (31%) | 131 (31%) |
| $3,000 or more ($156,000 or more) | 28 (25%) | 348 (19%) | 69 (16%) |
| How family is getting on financially (FS) | | | |
| Prosperous/Very comfortable | 39 (34%) | 649 (36%) | 122 (29%) |
| Comfortable/getting along | 74 (65%) | 1,166 (64%) | 298 (71%) |
| Poor/Very poor | 1 (0.9%) | 10 (0.5%) | 2 (0.5%) |

| Characteristic | Underweight<br>N = 114[1] | Normal<br>N = 1825[1] | Overweight<br>N = 422[1] |
|---|---|---|---|
| Hardship scale (FH) | 0.12 (0.48) | 0.15 (0.49) | 0.18 (0.50) |
| Parental school completion (P1E) | 86 (75%) | 1,437 (79%) | 303 (72%) |
| Parental school completion (P2E) | 76 (67%) | 1,225 (67%) | 267 (63%) |
| Parental Body Mass Index (BMI1) | 23.9 (4.7) | 25.4 (5.3) | 27.8 (5.7) |
| Parental Body Mass Index (BMI2) | 26.3 (5.0) | 27.1 (3.7) | 29.1 (4.7) |
| Frequency of feeling rushed (RP1) | | | |
| Always/Often | 70 (61%) | 1,101 (60%) | 256 (61%) |
| Sometimes | 35 (31%) | 594 (33%) | 126 (30%) |
| Rarely/Never | 9 (7.9%) | 130 (7.1%) | 40 (9.5%) |
| K-6 Depression scale summed score (DP1) | 8.89 (2.76) | 8.73 (2.99) | 8.93 (3.24) |
| Serves of fruit and vegetables (FV) | 2.32 (1.32) | 2.48 (1.48) | 2.50 (1.56) |
| Serves of high fat food (inc. whole milk) (HF) | 3.18 (1.66) | 3.58 (1.68) | 3.54 (1.86) |
| Serves of high sugar drinks (HSD) | 1.11 (1.07) | 1.06 (1.13) | 1.19 (1.28) |
| Poor sleep quality (SL)[a] | 45 (39%) | 601 (33%) | 135 (32%) |
| Wake up in the morning (Time) (SLD) | 638 (36) | 640 (47) | 636 (39) |
| Outdoor environment quality (OD) | 1.81 (0.52) | 1.77 (0.50) | 1.86 (0.50) |
| Child regularly spoken to in a language other than | 25 (22%) | 273 (15%) | 86 (20%) |
| No. weeks of gestation (GW) | 38.52 (5.77) | 38.83 (4.16) | 39.14 (2.83) |
| Birth weight z-score (BWZ) | -0.38 (0.91) | 0.01 (1.04) | 0.26 (1.04) |

[1]n (%); Mean (SD); [a]Sleep problems > 0

#### Table S7. Birth cohort boys aged 8 to 9

| Characteristic | Underweight<br>N = 50[1] | Normal<br>N = 838[1] | Overweight<br>N = 209[1] |
|---|---|---|---|
| Female | 0 (0%) | 0 (0%) | 0 (0%) |
| BMI z-score (BMI) | -1.84 (0.60) | 0.18 (0.60) | 1.68 (0.35) |
| socio-economic position (SE) | 0.42 (0.93) | 0.35 (0.90) | -0.02 (0.92) |
| Child's choice to spend free time (FTA) | | | |
| Active | 11 (22%) | 249 (30%) | 41 (20%) |
| Active and inactive | 27 (54%) | 362 (43%) | 88 (42%) |
| Inactive | 12 (24%) | 227 (27%) | 80 (38%) |
| Total No. of TV minutes for an average week (TV) | 12 (7) | 13 (8) | 15 (8) |
| Total No. of electronic game minutes for an average | 6.0 (6.0) | 5.9 (5.3) | 6.9 (6.7) |
| SDQ Emotional symptoms scale (EM) | 2.04 (1.89) | 1.48 (1.67) | 1.71 (1.86) |
| SDQ Conduct problems scale (CD) | 1.12 (1.14) | 1.23 (1.39) | 1.49 (1.42) |
| Weekly household income (annual) (INC) | | | |
| $0-$999 ($0-$51,999) | 2 (4.0%) | 42 (5.0%) | 29 (14%) |
| $1,000-$1,999 ($52,000-$103,999) | 18 (36%) | 259 (31%) | 68 (33%) |

| Characteristic | Underweight N = 50[1] | Normal N = 838[1] | Overweight N = 209[1] |
|---|---|---|---|
| $2,000-$2,999 ($104,000-$155,999) | 17 (34%) | 279 (33%) | 74 (35%) |
| $3,000 or more ($156,000 or more) | 13 (26%) | 258 (31%) | 38 (18%) |
| How family is getting on financially (FS) | | | |
| Prosperous/Very comfortable | 14 (28%) | 292 (35%) | 58 (28%) |
| Comfortable/getting along | 36 (72%) | 539 (64%) | 148 (71%) |
| Poor/Very poor | 0 (0%) | 7 (0.8%) | 3 (1.4%) |
| Hardship scale (FH) | 0.14 (0.35) | 0.13 (0.47) | 0.20 (0.58) |
| Parental school completion (P1E) | 38 (76%) | 664 (79%) | 149 (71%) |
| Parental school completion (P2E) | 32 (64%) | 582 (69%) | 131 (63%) |
| Parental Body Mass Index (BMI1) | 24.6 (5.3) | 25.5 (5.2) | 29.3 (6.2) |
| Parental Body Mass Index (BMI2) | 25.8 (3.3) | 27.3 (4.0) | 29.3 (4.6) |
| Frequency of feeling rushed (RP1) | | | |
| Always/Often | 32 (64%) | 510 (61%) | 108 (52%) |
| Sometimes | 16 (32%) | 277 (33%) | 79 (38%) |
| Rarely/Never | 2 (4.0%) | 51 (6.1%) | 22 (11%) |
| K-6 Depression scale summed score (DP1) | 8.06 (2.25) | 8.40 (2.89) | 9.11 (3.75) |
| Frequency ate fruit and vegetables (FV) | 3.34 (1.24) | 3.43 (1.41) | 3.09 (1.27) |
| Frequency ate high fat food (inc. whole milk) (HF) | 3.22 (1.39) | 3.29 (1.50) | 3.14 (1.52) |
| Frequency drank high sugar drinks (HSD) | 1.18 (1.06) | 0.96 (1.02) | 0.99 (1.02) |
| Poor sleep quality (SL)[a] | 17 (34%) | 232 (28%) | 60 (29%) |
| Wake up in the morning (Time) (SLD) | 610 (45) | 616 (38) | 603 (44) |
| Child regularly spoken to in a language other than | 10 (20%) | 116 (14%) | 38 (18%) |
| No. weeks of gestation (GW) | 38.28 (6.29) | 39.01 (2.75) | 39.15 (3.40) |
| Birth weight z-score (BWZ) | -0.37 (0.96) | 0.04 (1.02) | 0.25 (1.11) |

[1]n (%); Mean (SD); [a]Sleep problems > 0

#### Table S8. Birth cohort girls aged 8 to 9

| Characteristic | Underweight N = 57[1] | Normal N = 763[1] | Overweight N = 218[1] |
|---|---|---|---|
| Female | 57 (100%) | 763 (100%) | 218 (100%) |
| BMI z-score (BMI) | -1.87 (0.75) | 0.08 (0.57) | 1.55 (0.39) |
| Socio-economic position (SE) | 0.28 (1.04) | 0.30 (0.94) | 0.14 (0.84) |
| Child's choice to spend free time (FTA) | | | |
| Active | 18 (32%) | 181 (24%) | 45 (21%) |
| Active and inactive | 28 (49%) | 388 (51%) | 111 (51%) |
| Inactive | 11 (19%) | 193 (25%) | 62 (28%) |

| Characteristic | Underweight N = 57[1] | Normal N = 763[1] | Overweight N = 218[1] |
|---|---|---|---|
| Total No. of TV minutes for an average week (TV) | 13 (6) | 13 (8) | 13 (7) |
| Total No. of electronic game minutes for an average | 4.6 (4.5) | 4.0 (4.1) | 3.8 (3.0) |
| SDQ Emotional symptoms scale (EM) | 2.07 (2.09) | 1.76 (1.84) | 1.92 (1.87) |
| SDQ Conduct problems scale (CD) | 0.89 (1.14) | 0.92 (1.18) | 1.12 (1.44) |
| Weekly household income (annual) (INC) | | | |
| $0-$999 ($0-$51,999) | 8 (14%) | 44 (5.8%) | 17 (7.8%) |
| $1,000-$1,999 ($52,000-$103,999) | 16 (28%) | 257 (34%) | 65 (30%) |
| $2,000-$2,999 ($104,000-$155,999) | 20 (35%) | 261 (34%) | 80 (37%) |
| $3,000 or more ($156,000 or more) | 13 (23%) | 201 (26%) | 56 (26%) |
| How family is getting on financially (FS) | | | |
| Prosperous/Very comfortable | 11 (19%) | 241 (32%) | 55 (25%) |
| Comfortable/getting along | 46 (81%) | 518 (68%) | 160 (73%) |
| Poor/Very poor | 0 (0%) | 4 (0.5%) | 3 (1.4%) |
| Hardship scale (FH) | 0.16 (0.53) | 0.14 (0.54) | 0.23 (0.53) |
| Parental school completion (P1E) | 40 (70%) | 617 (81%) | 153 (70%) |
| Parental school completion (P2E) | 36 (63%) | 506 (66%) | 134 (61%) |
| Parental Body Mass Index (BMI1) | 23.9 (4.6) | 25.4 (5.0) | 28.3 (5.8) |
| Parental Body Mass Index (BMI2) | 25.6 (3.3) | 27.2 (3.9) | 29.5 (4.9) |
| Frequency of feeling rushed (RP1) | | | |
| Always/Often | 33 (58%) | 482 (63%) | 134 (61%) |
| Sometimes | 21 (37%) | 225 (29%) | 70 (32%) |
| Rarely/Never | 3 (5.3%) | 56 (7.3%) | 14 (6.4%) |
| K-6 Depression scale summed score (DP1) | 9.02 (2.91) | 8.44 (2.76) | 8.61 (2.87) |
| Frequency ate fruit and vegetables (FV) | 3.05 (1.26) | 3.39 (1.35) | 3.46 (1.46) |
| Frequency ate high fat food (inc. whole milk) (HF) | 3.18 (1.44) | 3.19 (1.34) | 3.11 (1.56) |
| Frequency drank high sugar drinks (HSD) | 0.82 (1.07) | 0.92 (1.01) | 1.09 (1.11) |
| Poor sleep quality (SL)[a] | 16 (28%) | 231 (30%) | 56 (26%) |
| Wake up in the morning (Time) (SLD) | 613 (42) | 621 (37) | 618 (41) |
| Child regularly spoken to in a language other than | 13 (23%) | 135 (18%) | 31 (14%) |
| No. weeks of gestation (GW) | 38.19 (5.80) | 38.82 (4.62) | 38.87 (3.49) |
| Birth weight z-score (BWZ) | -0.45 (1.01) | 0.04 (1.06) | 0.16 (1.08) |

[1]n (%); Mean (SD); [a]Sleep problems > 0

[2] Wanchuang Z, Ngoc LCN, Sally C. Structure Learning for Hybrid Bayesian Networks. arXiV, https://arxiv.org/abs/2206.01356
